# Targeting Lyn Kinase in Chorea-Acanthocytosis: A Translational Treatment Approach in an Ultra-Rare Disease

**DOI:** 10.1101/2021.03.08.21252937

**Authors:** Kevin Peikert, Hannes Glaß, Enrica Federti, Alessandro Matte, Lisann Pelzl, Katja Akgün, Tjalf Ziemssen, Rainer Ordemann, Florian Lang, The Network for translational research for Neuroacanthocytosis Patients, Lucia De Franceschi, Andreas Hermann

## Abstract

**Background:** Chorea-acanthocytosis (ChAc) is a neurodegenerative disease caused by mutations in the *VPS13A* gene. It is characterized by several neurological symptoms and the appearance of acanthocytes. Elevated tyrosine kinase Lyn activity has been recently identified as one of the key pathophysiological mechanisms and therefore represents a promising drug target.

**Methods:** We evaluated an individual off-label treatment with the FDA-approved tyrosine kinase inhibitor dasatinib (100 mg/d, 25.8-50.4 weeks) of three ChAc patients. Alongside with a thorough safety monitoring, we assessed motor and non-motor scales (e.g. MDS-UPDRS, UHDRS, quality of life) as well as routine and experimental laboratory parameters (e.g. serum neurofilament, Lyn kinase activity, actin cytoskeleton in red blood cells).

**Results:** Dasatinib appeared to be reasonably safe. The clinical parameters remained stable without significant improvement or deterioration. Regain of deep tendon reflexes was observed in one patient. Creatine kinase, serum neurofilament levels and acanthocyte count did not reveal consistent effects. However, reduction of initially elevated Lyn kinase activity and accumulated autophagy markers as well as partial restoration of actin cytoskeleton was found in red blood cells.

**Discussion:** We report on the first treatment approach with disease-modifying intention in ChAc. The experimental parameters indicate target engagement in red blood cells, while clinical effects on the central nervous system could not be proven within a rather short treatment time. Limited knowledge on the natural history of ChAc and the lack of appropriate biomarkers remain major barriers for “clinical trial readiness”. Here, we suggest a panel of outcome parameters for future clinical trials in ChAc.

## INTRODUCTION

Chorea-acanthocytosis (ChAc) is a rare neurodegenerative disease of the early adulthood which is characterized by a large spectrum of neurological symptoms and the presence of acanthocytes ^1-4^. The autosomal-recessive condition is caused by mutations in the *VPS13A* gene leading to loss of function of the respective encoded protein “chorein” ^5-8^. A disease-modifying therapy is not available yet. Hence, treatment options of the devastating disease remains purely symptomatic ^9^ even though it causes considerable morbidity, markedly reduced life-span and severely affects self-determined living.

The clinical phenotype of ChAc is highly heterogeneous. As patients often present with movement disorders like chorea, Parkinsonism and/or dystonia ^2, 10, 11^, ChAc belongs to the group of Huntington’s disease (HD) phenocopies ^12^. Furthermore, dysarthria and dysphagia, peripheral neuropathy, epilepsy or cognitive impairment may occur, while tongue and lip biting, self-mutilating behavior, feeding dystonia or head drops are more specific signs of ChAc ^1, 2, 10, 11^. Red blood cell (RBC) acanthocytosis, elevated creatine kinase (CK) and serum neurofilament (sNfL) levels are common laboratory findings ^1, 2, 10, 13^. In correlation with the clinical manifestations, loss of striatal medium spiny neurons and distinct cortical neurodegeneration represent the main histopathological characteristics ^14, 15^. Epidemiological estimations suggest a prevalence of around 1000-5000 cases worldwide ^1^.

While there is growing evidence that members of the VPS13 protein family are involved in the non-vesicular transport of phospholipids ^16-19^, the precise function of these proteins in humans remains incompletely understood. So far, VPS13A has been implicated in a variety of important cell processes, e.g., regulation of cytoskeletal architecture, exocytosis, autophagy, Ca^2+^ homeostasis and therefore overall cell survival ^4, 20-24^. Hence, lack of functional VPS13A leads to impaired cellular homeostasis particularly resulting in acanthocytosis and neurodegeneration. We recently identified two mechanisms that are considered to be key drivers of ChAc pathophysiology due to VPS13A deficiency: decreased phosphoinositide-3-kinase (PI3K) signaling and increased activity of Src family tyrosine kinase Lyn (for review, see ^4^).

In previous studies, we found that ChAc RBCs and induced pluripotent stem cell (iPSC) derived nerve cells are characterized by accumulation of active Lyn: Hyperactive Lyn kinase hyperphosphorylates membrane proteins in RBCs, e.g., band 3, which is involved in anchoring the membrane to the cytoskeletal network. This results in mechanical instability of the membrane ^21, 25^. Accumulation of active Lyn was also found to be related to impairment of autophagy in erythrocytes ^21^. Consistent with that, perturbations of autophagy processes have been reported in *in vitro* cell models defective for VPS13A ^17, 26^. In a previous study, we could show that *in vitro* treatment of ChAc RBCs with Src family kinase inhibitors reduces Lyn kinase activity, improves autophagy and restores the morphological phenotype ^21^. Moreover, Src family kinase inhibition also attenuated pathologically enhanced synaptic transmission in striatal medium spiny neurons derived from patient specific iPSCs ^27^. These findings strongly implicate that Lyn kinase should be considered as a promising potential druggable target in ChAc. FDA-approved specific inhibitors of Src family kinases with a reasonable benefit-risk profile such as the tyrosine kinase inhibitor (TKI) dasatinib were successfully established in the treatment of chronic myeloid leukemia ^28^. Dasatinib was previously shown to cross the blood brain barrier^29^. Therefore, they are ideal candidates for “repurposing” strategies in this context. TKIs are currently being evaluated in a variety of other neurodegenerative diseases, e.g. Parkinson’s disease (nilotinib), Alzheimer’s disease (e.g., nilotinib, saracatinib), or Amyotrophic Lateral Sclerosis (masitinib), each with a different pathophysiological rationale ^28, 30-34^.

We here evaluate a translational off-label treatment with dasatinib (100 mg/d) in three ChAc patients. To our knowledge, this is the first implemented potentially disease modifying approach in this ultra-rare disease. Alongside with a thorough safety monitoring, we regularly performed assessments of treatment efficacy. Therefore, we designed a novel ChAc-related panel of read out parameters also challenging current concepts of disease markers.

## METHODS

### Patients and dasatinib treatment

We treated three ChAc patients (P1-3) off-label with the FDA-approved tyrosine kinase inhibitor (TKI) dasatinib (DRKS00023177). Patient characteristics are shown in Table 1 and reflected the variety of ChAc phenotypes. Diagnosis was based on clinical manifestations, the absence of chorein in Western blot and genetic testing ^35^. All patients gave their informed consent for off-label treatment with dasatinib, including the risk of possible life-threatening adverse reactions, as well as for video documentation and publication of the data. Patients and healthy control blood donors were enrolled in ongoing studies on the pathogenesis and natural history of neurodegenerative diseases approved by the institutional review board of the Technische Universität Dresden, Germany (EK 45022009, EK 78022015). The standard dose of 100 mg dasatinib per day was administered orally.

**Table 1.**
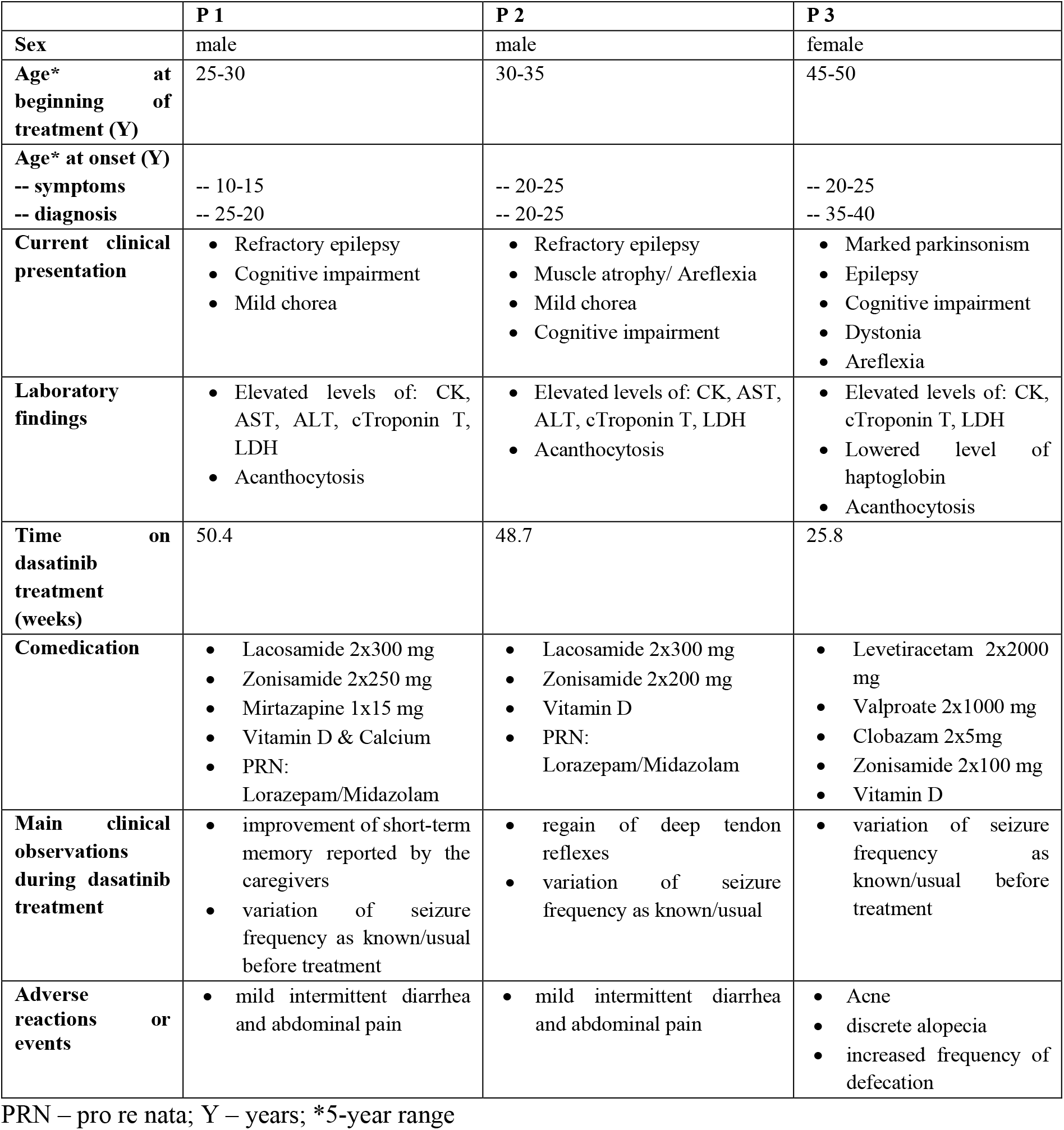
Demographic characteristics, main clinical observations and adverse reactions/events in chorea-acanthocytosis patients treated with dasatinib.

### Evaluation of dasatinib treatment

Evaluation of the off-label treatment primarily included monitoring of potential adverse reactions or events, as well as outcome assessments in the context of routine care adjusted to the clinical phenotype (UHDRS, MDS-UPDRS, seizure frequency, blood acanthocyte and CK level). Evaluation was performed initially every 2 weeks; after 8 weeks of treatment monthly; and after 6 months of treatment every two months. At every visit, each patient was assessed with a clinical examination and medical history (including the caregivers’ observations), the Unified Huntington’s Disease Rating Scale Total Motor Score (UHDRS-TMS), Total Functional Capacity (UHDRS-TFC) and Functional Assessment Scale (UHDRS-FA), the Movement Disorders Society Unified Parkinson Disease Rating Scale (MDS-UPDRS) parts I-III and the Clinical Global Impression (GCI) scale by a specialist experienced in the care of patients with ChAc and other movement disorders. Patients were asked to complete the McGill Quality of Life Single Item Scale (McGill-QoL; range 0-10) and the Schedule for the Evaluation of Individual Quality of Life-Direct Weighting (SEIQoL) questionnaires. Routine electroencephalography was performed. Seizure frequency was defined as number of seizures within the preceding month. At baseline, after two months, and as indicated, patients underwent electrocardiography, echocardiography, and abdominal ultrasound for safety monitoring. Routine laboratory chemistry/haematology tests were also performed on blood samples obtained by venipuncture.

### Serum neurofilament light chain quantification

Serum samples were stored at −20°C directly after collection since neurofilament light chain is stable during freezing process ^36, 37^. sNfl measurement was performed using the Advantage NF-Light singleplex Kit and prepared as defined in the manufacturer’s instructions (Quartered, Lexington, MA, Datasheet Quanterix: Simoa™ NF-Light® Advantage Kit) as previously described ^38, 39^ with the single molecule array (SIMOA) analysis. Both the mean intra-assay coefficient of variation of duplicates and the mean inter-assay coefficient of variation was <10%.

### Immunoblot analysis

Additional EDTA-blood samples (ChAc patients and healthy control donors) were shipped to Verona, Italy at 4°C and processed immediately after arrival. Lyn activity was determined by Western blot analysis using anti-phospho-Lyn (Y396) antibody as described previously ^25^. We evaluate the amount of ULK1 and p62, known markers of autophagy in RBCs, as reported previously ^21, 40, 41^.

### Immunofluorescence

Additional EDTA-blood samples (ChAc patients and healthy control donors) were shipped to Tübingen, Germany at 4°C and processed immediately after arrival. The erythrocytes were stained with anti-ß-Actin-FITC-conjugated antibody (1:50; biorbyt) and Phalloidin-eFluor660 (1:100, eBioscience) to detect filamentous actin (F-actin) as described previously ^24, 42^. Confocal microscopy was performed with a Zeiss LSM 5 EXCITER confocal laser-scanning module (Carl Zeiss). The images were analyzed with the software of the instrument.

### Osmotic fragility of red blood cells

In previous reports, we showed that red cell osmotic fragility is increased in patients with ChAc compared to healthy controls ^21, 25, 43, 44^. We evaluated osmotic fragility in EDTA blood using a single osmotic point at 158 mOsm. Erythrocytes from healthy controls were always analyzed in the same experiments with ChAc RBCs.

### Statistical analysis

Data were analyzed with GraphPad Prism 5 software. Statistical analysis was made by analysis of variance (ANOVA). p < 0.05 was considered as statistically significant.

## RESULTS

### Dasatinib treatment was safe

Overall, dasatinib treatment was safe in all three patients (Table 1). No severe adverse reactions or events were reported. Due to discrete acne and alopecia as well as increased defecation frequency, P3 and her caregivers asked for discontinuation of treatment after 25.8 weeks. Otherwise, P1 and P2 reported on irregular defecation including episodes of mild diarrhea and abdominal pain, but they both stayed on dasatinib treatment for 48.7 and 50.4 weeks, respectively.

We observed a reversible decrease of RBC level and hematocrit, but no manifest anemia in all three patients in the first weeks after treatment initiation (Figure 1 A, B). No major change in haptoglobin, a marker of hemolysis, was detected during dasatinib administration (Figure 1 C); Mean corpuscular volume (MCV), mean corpuscular hemoglobin (MCH) and red blood cell distribution width (RDW) increased slightly under medication especially in P3 (Figure 1 D-F). In the first week of treatment, we observed a transient thrombocytopenia (minimum 107 GPt/l in P1). No relevant neutropenia <1.5 GPt/l nor thrombopenia <100 GPt/l occurred. After discontinuation of dasatinib, there was a drop in hemoglobin and haptoglobin, suggesting a transient hemolytic episode in P1 and P2 (Figure 1 C). Taken together, these recorded mild symptoms and laboratory abnormalities are known adverse reactions/events of dasatinib, which did not require to discontinue the treatment ^45^. Dasatinib was withdrawn after 50.4 (P1) and 48.7 (P2) weeks for re-evaluation purposes as predetermined with the health insurance.

**Figure 1.**
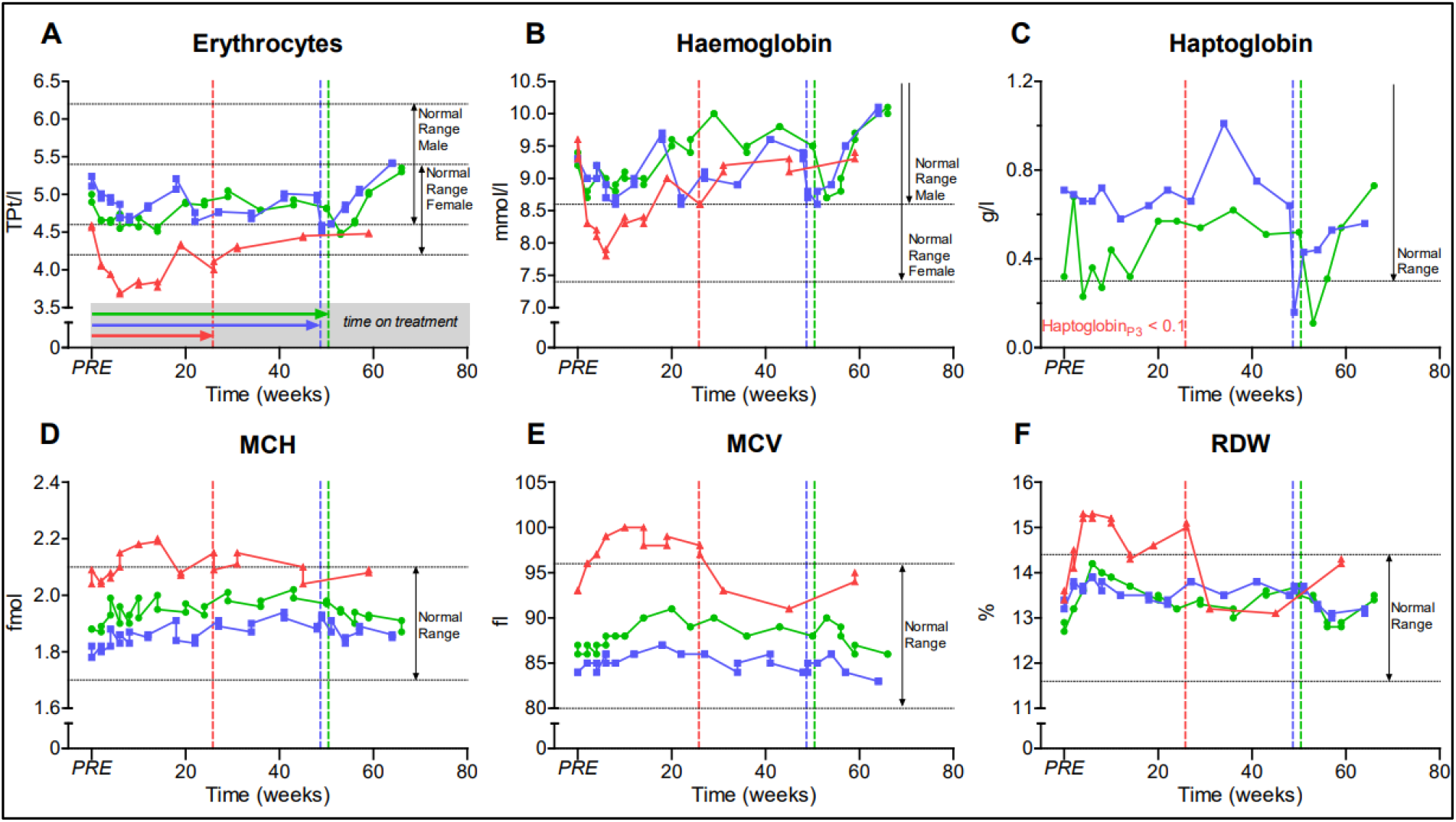
Graphs represent hematology parameters. Patient 1-3 (P1-3), PRE represents baseline value before initiation of dasatinib treatment, dotted vertical lines represent individual end of treatment. Mean corpuscular volume (MCV); Mean corpuscular hemoglobin (MCH); Red blood cell distribution width (RDW).

### Disease progression remained stable during dasatinib treatment in ChAc patients

During dasatinib treatment neither significant clinical improvement nor deterioration of neurological manifestations was observed in the three ChAc patients. Caregivers of P1 reported an improvement of short-term memory. In P2, who was initially noted to have areflexia, the re-appearance of patellar (after 2 weeks) and biceps (after 4 weeks) deep tendon reflexes was a striking observation (Table 1).

Quality of life improved for a short time period after initiation of treatment (Figure 2 A, B) but dropped to baseline level before treatment was withdrawn, most likely representing a placebo effect. Seizure frequency (Figure 2 C) fluctuated widely and was associated with physical and mental stress in P1 and P2. Unified Huntington’s Disease Rating-Scale (UHDRS) and Movement Disorders Society Unified Parkinson Disease Rating Scale (MDS-UPDRS) scores remained stable without clinically relevant progression (Figure 2 D-I). Electroencephalograms at baseline and under treatment revealed no significant changes during treatment.

**Figure 2.**
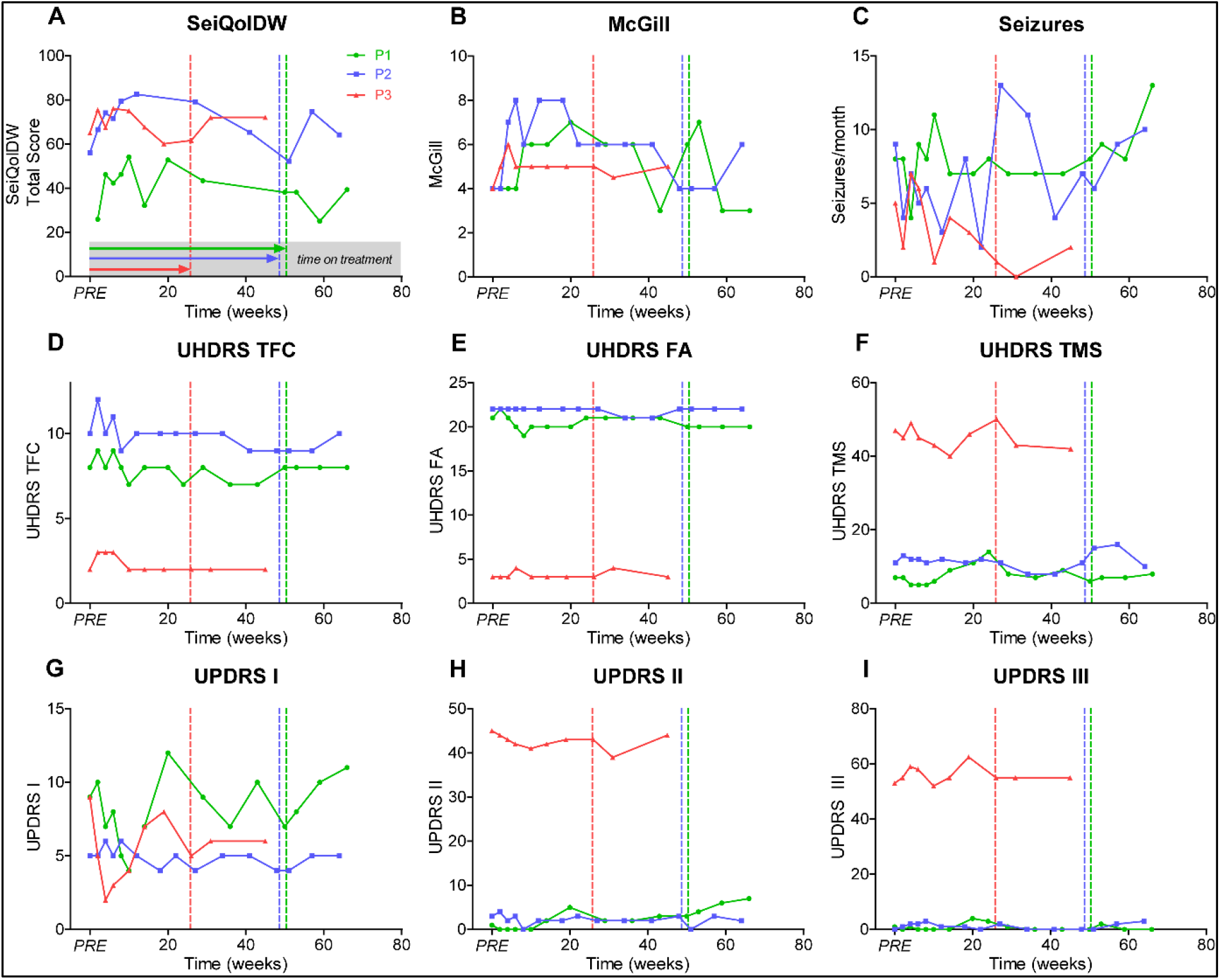
Graphs represent **A)** Schedule for the Evaluation of Individual Quality of Life-Direct Weighting (SEIQoL); **B)** McGill Quality of Life Single Item Scale (McGill); **C)** Seizure frequency per month; **D)** Unified Huntington’s Disease Rating-Scale Total Functional Capacity (UHDRS TFC); **E)** UHDRS Functional Assessment Scale (UHDRS FA); **F)** UHDRS Total Motor Scale (UHDRS TMS); **G)** Movement Disorders Society Unified Parkinson’s Disease Rating Scale part I (UPDRS I); **H)** MDS-UPDRS part II (UPDRS II) and **I)** MDS-UPDRS part III (UHDRS III). Patient 1-3 (P1-3), PRE represents baseline value before initiation of dasatinib treatment, dotted vertical lines represent individual end of treatment.

### Identification of robust biomarkers is still an unmet need in chorea-acanthocytosis

We next investigated whether dasatinib treatment alters current biomarker candidates of the disease. Of those, the amount of acanthocytes showed severe variations. The acanthocyte level of P1 (range 21.4 %-42.0 %, at baseline 35.2 %) and P2 (range 19.4 %-44.6%, at baseline 24.2 %) varied markedly during the treatment without a clear trend. P3 (range 12.7 %-28.9 %, at baseline 13.5 %) showed a trend to increased acanthocyte levels under treatment and subsequent decrease after dasatinib withdrawal (Figure 3 A). However, limitations of the accuracy of the acanthocyte testing (manual counting) and pre-analytical variations (transport time to the laboratory etc.) have to be taken into account just like the intraindividual variations on the amount of acanthocytes in ChAc.

**Figure 3.**
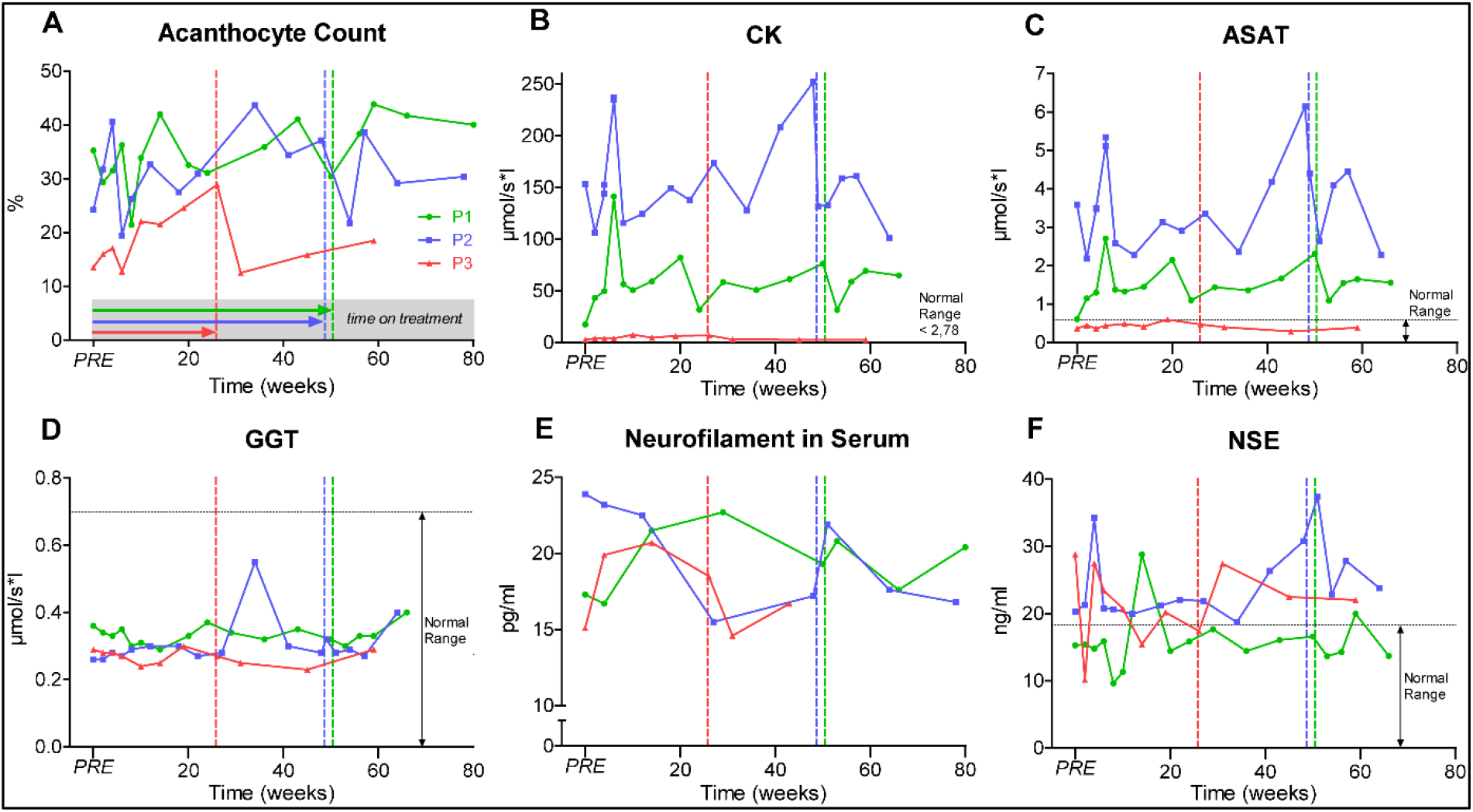
Graphs represent ChAc biomarkers. Patient 1-3 (P1-3), PRE represents baseline value before initiation of dasatinib treatment, dotted vertical lines represent individual end of treatment. Alanine aminotransferase (ALAT); Gamma-glutamyltransferase (GGT).

Elevated creatine kinase (CK) persisted in P1 and P2 throughout the study but increased slightly in P3 (Figure 3 B). As CK markedly fluctuated due to athletic activity and tonic-clonic seizure activity, it was not a robust clinical marker of clinical severity in ChAc. Alanine aminotransferase (ALAT) and aspartate aminotransferase (ASAT) levels correlated with CK while gamma-glutamyltransferase (GGT) and bilirubin remained rather stable within the physiological range suggesting a most likely muscular origin of the increased ASAT and ALAT activities (Figure 3 C-D).

Neurofilament light chain (Nfl) was here evaluated as a biomarker for neuroaxonal damage^13, 46^. As shown in Figure 3 E, Nfl varied through the observation period (16.7-22.7 pg/ml in P1; 15.5– 23.9 pg/ml in P2; 14.6-20.7 pg/ml in P3). While Nfl concentrations showed a trend to decrease under treatment in P2, the levels of P1 and P3 increased. The concentration of other peripheral markers for neuronal destruction, S100 and neuron-specific enolase (NSE), were subject to fluctuation without an evident relevant trend (Figure 3 F).

### Dasatinib treatment prevented accumulation of Lyn and autophagy-related proteins and improved features of ChAc RBCs

In RBCs from all three ChAc patients, dasatinib markedly reduced the amount of initially increased active Lyn (phosphor-Lyn) compared to healthy controls (Figure 4 A-C). Active Lyn abundance progressively increased after dasatinib was withdrawn. It is of note that we observed in P1 and P2 (Figure 4 A-B) a slight and temporary increase in Lyn activation after 12 and 14 weeks of treatment, respectively, to values similar to those of controls but still lower compared to baseline.

**Figure 4.**
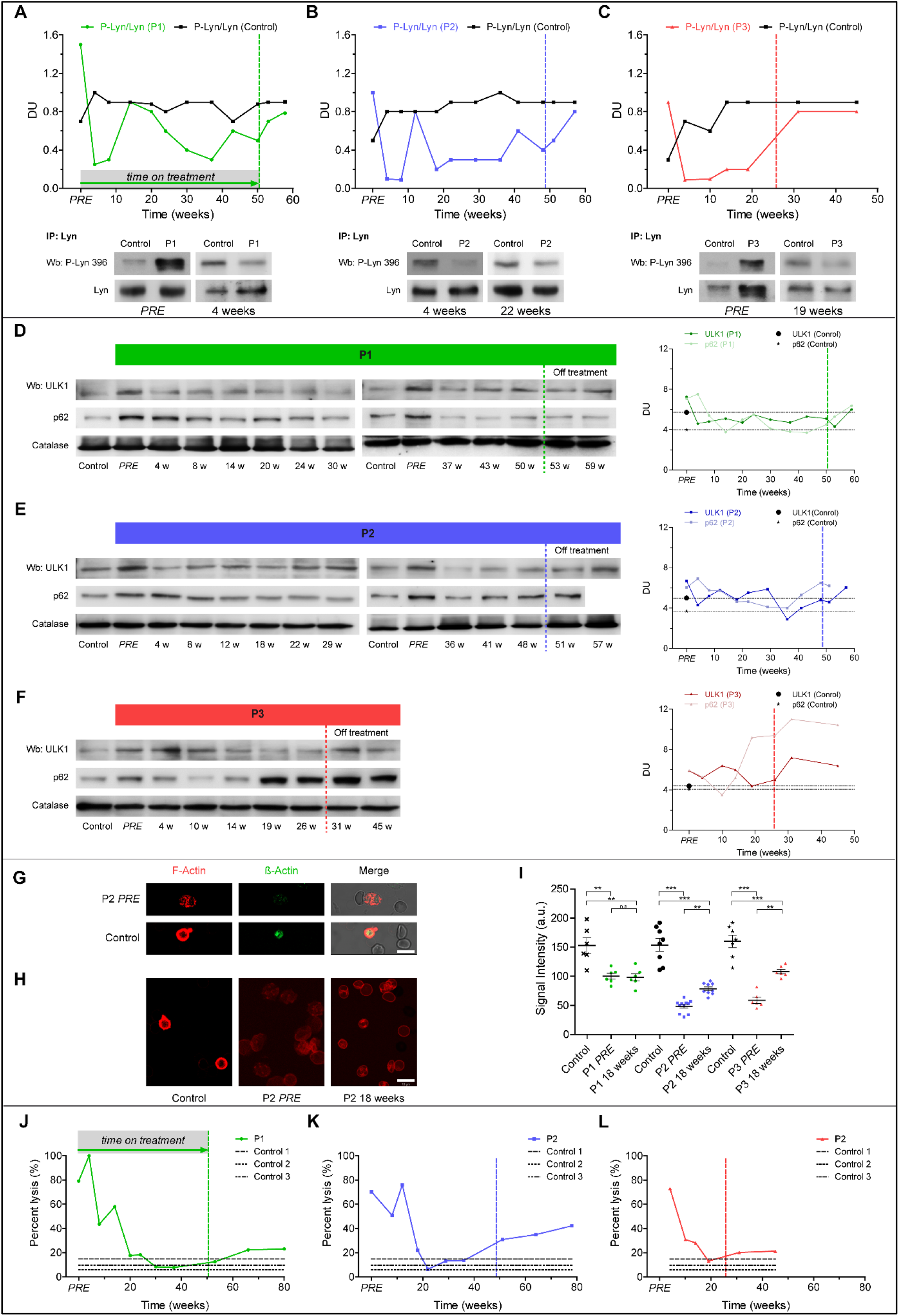
Graphs represent red cell features in ChAc patients. **A-C)** Total Lyn was immunoprecipitated from blood samples and detected with antibody against active Lyn (phospho-Lyn 396) or total Lyn (Wb: Western blot). Initially overactive Lyn was suppressed by dasatinib treatment in all three patients. Lyn activity increased after dasatinib withdrawal. **D-F)** Western blot (Wb) analysis of Ulk1 and p62 in red blood cells (RBCs) and densitometric analyses of the immunoblot bands. **G)** Confocal microscopy of microfilament actin-fluorescence and transmission light microscopy of RBCs from patient 2 (P2) and healthy control. Anti-β-Actin-FITC-conjugated antibody stained total actin and Phalloidin stained filamentous actin (F-actin). **H)** Phalloidin was used for F-actin staining. Scale bar: 10 μm. **I)** F-actin staining intensity in P1-P3. Shown are the mean values of intensity ± SEM from n=3 independent experiments. One-way-ANOVA/Bonferroni’s multiple comparison test (p<0.05). **J-L)** Osmotic fragility of RBCs is shown as percentage of RBC lysis at a single osmotic point at 158 mOsm in P1-P3. Patient 1-3 (P1-3), PRE: represents baseline value before initiation of dasatinib treatment, dotted vertical lines represent individual end of treatment.

Autophagy initiator ULK1 markedly decreased after 4 weeks of treatment in all patients (Figure 4 D-F) and increased after dasatinib withdrawal. The accumulation of the late phase autophagy marker p62 markedly decreased between 14 and 24 weeks of treatment in P1 (Figure 4 D) and between 12 and 22 weeks of dasatinib in P2 (Figure 4 E). In contrast, p62 accumulation in P3 RBCs was only temporarily reduced versus baseline (Figure 4 F) and re-accumulation along with active Lyn before the schedule interruption of dasatinib treatment was consistent with possibly uncompliant dosing.

We revealed a severe loss of cortical actin in RBCs from P2 and P3 as compared to healthy controls at baseline, whereas the cortical actin network was less impaired in P1 (Figure 4 G, H). Consistent with the findings described above, we observed a slight but significant increase of F-actin staining in RBCs from P2 and P3 (Figure 4 H, I) associated with improvement/normalization of osmotic fragility (Figure 4 J-L). After withdrawal of dasatinib, osmotic fragility again progressively deteriorated (Figure 4 J-L).

## DISCUSSION

Chorea-acanthocytosis is a devastating neurodegenerative multi-system disorder without any established disease modifying treatment. In our previous work we could show that elevated Lyn kinase activity represents a pathophysiological hallmark of the disease ^21, 25, 27^. FDA-approved TKIs like dasatinib with specificity to Lyn kinase are therefore promising candidates for a potential disease modifying treatment. Herein, we report on three phenotypically different ChAc patients who were prescribed dasatinib and monitored for potential adverse reactions or events ^45^ and therapy response.

Dasatinib successfully engaged the target Lyn kinase with subsequent normalization of ChAc RBC features, but neither showed positive nor negative effects on central nervous system parameters in the three ChAc patients. Oral administration of 100 mg dasatinib once a day appeared to be reasonably safe in subjects with ChAc. Only rather mild adverse reactions including gastrointestinal and cutaneous problems occurred and hematological adverse reactions were transient and well tolerated.

Dasatinib sufficiently inhibited Lyn kinase at least in RBCs. In all patients, the phospho-Lyn/total Lyn ratio in erythrocytes was pathologically increased at baseline consistent with our previous findings ^21, 25^. The ratio markedly decreased under dasatinib treatment. Moreover, Lyn kinase inhibition improved autophagy as supported by the reduction of accumulated autophagy related proteins ^21^ and led to a rearrangement of the initially severely impaired cortical filamentous actin network. Together with the reduction of the osmotic fragility these findings implicate improved mechanical properties of ChAc RBC membranes in dasatinib-treated subjects.

On the other hand, clinical motor and non-motor parameters could not show a clear treatment effect. Considering the rather slow progression rate of ChAc, however, absence of an immediate short-term improvement with such a disease modifying approach is not surprising. Since limited knowledge regarding the natural history and the heterogeneity of the disease course complicate interpretation of this data, it remains uncertain if the lack of evident clinical progression in all three patients can be attributed to positive treatment effects. Most notable was the regain of deep tendon reflexes in P2, which might point to a neuromuscular (peripheral) treatment response. sNfl levels in cerebrospinal fluid or in serum reflects neuroaxonal damage both of the central and the peripheral nervous system ^46^ and is known to be elevated in ChAc ^13^. Under dasatinib treatment, there was a trend to decreased sNfl concentration in the patient who regained deep tendon reflexes, however an increase in the other two patients. It is not clear if this course is due to the therapy or if it reflects intraindividual variations. Therefore, sNfl and other brain damage markers needs to be further established as biomarker for ChAc by long-term profiling of the individual levels.

One main finding of the study is that currently believed evident read-out parameters for ChAc (e.g., acanthocyte count, elevated CK, seizure frequency) which were hypothesized to show possible short time effects all failed to be useful in a stringent longitudinal follow-up of these patients. Particularly CK turned out to be subject of pronounced fluctuations due to athletic activity and bilateral convulsive seizures. Also, the amount of acanthocytes could not be used as parameter to evaluate the drug effect due to limitations of the accuracy of the acanthocyte testing and significant intraindividual variations on the amount of acanthocytes in ChAc. Thus, all these parameters are not as robust as expected which makes it - combined with the slow progression of the disease - currently difficult to perform clinical studies in this population of patients. With respect to the limited knowledge of the natural history, defining appropriate read-out parameters remains challenging. This is still, however, a classical translational roadblock in ultra-rare diseases in general. Larger observational studies such as worldwide case registers like the Neuroacanthocytosis database, a submodule of the European Huntington’s Disease Network (https://www.euro-hd.net/html/na/submodule), need to be further supported and enlarged in order to establish robust biomarkers and to enable “clinical trial readiness”. Our study represents an additional contribution to achieve this objective as we suggest a novel panel of outcome parameters for future clinical studies and for cohort characterization in ChAc.

Further open questions relate to the duration and time point of the treatment. It can be assumed that disease modification is more effective in the early stages of disease. However, all of our patients experienced first symptoms already 9 to 28 years prior to dasatinib therapy which might limit its effects. Considering the rather low progression rate of ChAc, longer treatment periods might be necessary to achieve.

In conclusion, this is the first report on a potentially disease modifying treatment approach in ChAc. Our observations during an off-label treatment suggest that dasatinib is safe and may successfully target pathologically accumulated Lyn kinase at least in RBCs. However, only a very small number of patients was treated over a rather short time period which are major limitations of this study. Nevertheless, our observations warrant moving forward with the evaluation of Lyn kinase inhibitor effects in larger studies. This study also reveals the need for further investigations of the natural history and robust biomarkers of the disease.

## Data Availability

Data is available upon request.

## Acknowledgements

The authors thank all patients and their families for giving consent for publication of the data and the healthy control subjects for participation in this study.

Expenses for the off-label dasatinib prescription were gratefully covered in part by the Center for Regenerative Therapies Dresden (CRTD, TU Dresden, Germany) and the public health care insurance of the patients. We are especially grateful to Glenn (†) and Ginger Irvine as founders of the Advocacy for Neuroacanthocytosis Patients (www.naadvocacy.org) and to Susan Wagner and Joy Willard-Williford as representatives of the NA Advocacy USA (www.naadvocacyusa.org).

Diagnostic Western blot analysis for chorein had been performed by G. Kwiatkowski and Dr. Benedikt Bader with the financial support of the Advocacy for Neuroacanthocytosis Patients and of the ERA-net E-Rare consortium EMINA (European Multidisciplinary Initiative on Neuroacanthocytosis; BMBF01GM1003) in the labs of Profs. Hans Kretzschmar/Armin Giese (Neuropathology) and Adrian Danek (Neurology) at Ludwig-Maximilians-Universität Munich, Germany.

K.P. was supported by the Else Kröner clinician scientist program and the MeDDrive program (TU Dresden, Germany) as well as by the Rostock Academy for Clinician Scientists (RACS) and the FORUN program (University of Rostock, Germany). A.H. was supported by the “Hermann und Lilly Schilling-Stiftung für medizinische Forschung im Stifterverband”.

## The Network for translational research for Neuroacanthocytosis Patients

**Enrico Bonifacio**, Center for Regenerative Therapies Dresden (CRTD), Technische Universität Dresden, Dresden, Germany

**Giel Bosman**, Department of Biochemistry, Radboud University Medical Center, Nijmegen, The Netherlands **Adrian Danek**, Neurologische Klinik, Klinikum der Universität, Ludwig-Maximilians-Universität München, Munich, Germany

**Peter Claus**, Institute of Neuroanatomy and Cell Biology, Hannover Medical School, Hannover, Germany and Center for Systems Neuroscience (ZSN) Hannover, Hannover, Germany

**Lucia de Franceschi**, Department of Medicine, University of Verona, Verona, Italy

**Jochen Guck**, Biotechnology Center, Dresden University of Technology, Dresden, Germany

**Andreas Hermann (Coordinator)**, Translational Neurodegeneration Section „Albrecht-Kossel”, Department of Neurology, University Medical Center Rostock, University of Rostock, Rostock, Germany

**Florian Lang**, Department of Physiology I, University of Tübingen, Tübingen, Germany

**Ulrich Salzer**, Department of Medical Biochemistry, Max F. Perutz Laboratories, Medical University of Vienna, Vienna, Austria

**Tjalf Ziemssen**, Department of Neurology, University Hospital Carl Gustav Carus, Technische Universität Dresden, Dresden, Germany

## Authors’ Roles

1. Research project: A. Conception, B. Organization, C. Execution;
2. Statistical Analysis: A. Design, B. Execution, C. Review and Critique;
3. Manuscript: A. Writing of the first draft, B. Review and Critique.

KP 1 A-C, 2A/B, 3 A

HG 1 B/C, 2A-C, 3 B

AM 1 C,

EF 1 C

LP 1 C, 2 B, 3B

KA 1 C, 2 B, 3B

TZ 3 B

RO 1 A/B, 3 B

FL 1 B, 3 B

LDF 1 B, 3 B

AH 1 A-C, 2A/B, 3 B

## Notes

**Funding sources for study:** This study was supported by the Centre for Regenerative Therapies Dresden (CRTD), the German Center for Neurodegenerative Diseases (DZNE), research site Dresden, the Helmholtz Virtual Institute (VH-VI-510), the Else Kröner Clinician Scientist Program (TU Dresden, Germany) and the Rostock Academy for Clinician Scientists (RACS, University of Rostock, Germany).

### Competing Interest Statement

The authors have declared no competing interest.

### Clinical Trial

DRKS00023177

### Funding Statement

This study was supported by the Centre for Regenerative Therapies Dresden (CRTD), the German Center for Neurodegenerative Diseases (DZNE), research site Dresden, the Helmholtz Virtual Institute (VH-VI-510), the Else Kroener Clinician Scientist Program (TU Dresden, Germany) and the Rostock Academy for Clinician Scientists (RACS, University of Rostock, Germany).

### Author Declarations

Patients and healthy control blood donors were enrolled in ongoing studies on the pathogenesis and natural history of neurodegenerative diseases approved by the institutional review board of the Technische Universitaet Dresden, Germany (EK 45022009, EK 78022015).

## References

1. Jung HH, Danek A, Walker RH. Neuroacanthocytosis syndromes. Orphanet journal of rare diseases 2011;6:68.

2. Velayos Baeza A, Dobson-Stone C, Rampoldi L, et al. Chorea-Acanthocytosis. In: Adam MP, Ardinger HH, Pagon RA, et al., eds. GeneReviews. Seattle (WA): University of Washington, Seattle, 1993.

3. Walker RH, Danek A. “Neuroacanthocytosis” - Overdue for a Taxonomic Update. Tremor Other Hyperkinet Mov (N Y) 2021;11:1.

4. Peikert K, Danek A, Hermann A. Current state of knowledge in Chorea-Acanthocytosis as core Neuroacanthocytosis syndrome. Eur J Med Genet 2018;61(11):699–705.

5. Danek A, Bader B, Velayos-Baeza A, Walker RH. Autosomal recessive transmission of chorea-acanthocytosis confirmed. Acta neuropathologica 2012;123(6):905–906.

6. Dobson-Stone C, Danek A, Rampoldi L, et al. Mutational spectrum of the CHAC gene in patients with chorea-acanthocytosis. European journal of human genetics : EJHG 2002;10(11):773–781.

7. Rampoldi L, Dobson-Stone C, Rubio JP, et al. A conserved sorting-associated protein is mutant in chorea-acanthocytosis. Nature Genetics 2001;28:119.

8. Ueno S-i, Maruki Y, Nakamura M, et al. The gene encoding a newly discovered protein, chorein, is mutated in chorea-acanthocytosis. Nature Genetics 2001;28:121.

9. Walker RH. Management of Neuroacanthocytosis Syndromes. Tremor and other hyperkinetic movements (New York, NY) 2015;5:346.

10. Rampoldi L, Danek A, Monaco AP. Clinical features and molecular bases of neuroacanthocytosis. Journal of molecular medicine (Berlin, Germany) 2002;80(8):475–491.

11. Estevez-Fraga C, Lopez-Sendon Moreno JL, Martinez-Castrillo JC. Phenomenology and disease progression of chorea-acanthocytosis patients in Spain. Parkinsonism & related disorders 2018;49:17–21.

12. Hermann A, Walker RH. Diagnosis and treatment of chorea syndromes. Current neurology and neuroscience reports 2015;15(2):514.

13. Peikert K, Akgün K, Beste C, et al. Neurofilament light chain in serum is significantly increased in chorea-acanthocytosis. Parkinsonism Relat Disord 2020;80:28–31.

14. Liu J, Heinsen H, Grinberg LT, et al. Subcortical neurodegeneration in chorea: Similarities and differences between chorea-acanthocytosis and Huntington’s disease. Parkinsonism & related disorders 2018;49:54–59.

15. Liu J, Heinsen H, Grinberg LT, et al. Pathoarchitectonics of the cerebral cortex in chorea-acanthocytosis and HD. Neuropathology and applied neurobiology 2018.

16. Gao M, Yang H. VPS13: A lipid transfer protein making contacts at multiple cellular locations. The Journal of cell biology 2018;217(10):3322–3324.

17. Kumar N, Leonzino M, Hancock-Cerutti W, et al. VPS13A and VPS13C are lipid transport proteins differentially localized at ER contact sites. The Journal of cell biology 2018;217(10):3625–3639.

18. Yeshaw WM, van der Zwaag M, Pinto F, et al. Human VPS13A is associated with multiple organelles and influences mitochondrial morphology and lipid droplet motility. eLife 2019;8.

19. Park JS, Neiman AM. XK is a partner for VPS13A: a molecular link between Chorea-Acanthocytosis and McLeod Syndrome. Mol Biol Cell 2020;31(22):2425–2436.

20. Lang F, Pelzl L, Schols L, et al. Neurons, Erythrocytes and Beyond -The Diverse Functions of Chorein. Neuro-Signals 2017;25(1):117–126.

21. Lupo F, Tibaldi E, Matte A, et al. A new molecular link between defective autophagy and erythroid abnormalities in chorea-acanthocytosis. Blood 2016;128(25):2976–2987.

22. Munoz-Braceras S, Calvo R, Escalante R. TipC and the chorea-acanthocytosis protein VPS13A regulate autophagy in Dictyostelium and human HeLa cells. Autophagy 2015;11(6):918–927.

23. Pelzl L, Elsir B, Sahu I, et al. Lithium Sensitivity of Store Operated Ca2+ Entry and Survival of Fibroblasts Isolated from Chorea-Acanthocytosis Patients. Cellular physiology and biochemistry : international journal of experimental cellular physiology, biochemistry, and pharmacology 2017;42(5):2066–2077.

24. Foller M, Hermann A, Gu S, et al. Chorein-sensitive polymerization of cortical actin and suicidal cell death in chorea-acanthocytosis. FASEB journal : official publication of the Federation of American Societies for Experimental Biology 2012;26(4):1526–1534.

25. De Franceschi L, Tomelleri C, Matte A, et al. Erythrocyte membrane changes of chorea-acanthocytosis are the result of altered Lyn kinase activity. Blood 2011;118(20):5652–5663.

26. Munoz-Braceras S, Tornero-Ecija AR, Vincent O, Escalante R. VPS13A is closely associated with mitochondria and is required for efficient lysosomal degradation. Dis Model Mech 2019;12(2).

27. Stanslowsky N, Reinhardt P, Glass H, et al. Neuronal Dysfunction in iPSC-Derived Medium Spiny Neurons from Chorea-Acanthocytosis Patients Is Reversed by Src Kinase Inhibition and F-Actin Stabilization. The Journal of neuroscience : the official journal of the Society for Neuroscience 2016;36(47):12027–12043.

28. Roskoski R, Jr. Src protein-tyrosine kinase structure, mechanism, and small molecule inhibitors. Pharmacological research 2015;94:9–25.

29. Porkka K, Koskenvesa P, Lundan T, et al. Dasatinib crosses the blood-brain barrier and is an efficient therapy for central nervous system Philadelphia chromosome-positive leukemia. Blood 2008;112(4):1005–1012.

30. Khairoalsindi OA, Abuzinadah AR. Maximizing the Survival of Amyotrophic Lateral Sclerosis Patients: Current Perspectives. Neurology research international 2018;2018:6534150.

31. Trias E, Ibarburu S, Barreto-Nunez R, et al. Post-paralysis tyrosine kinase inhibition with masitinib abrogates neuroinflammation and slows disease progression in inherited amyotrophic lateral sclerosis. Journal of neuroinflammation 2016;13(1):177.

32. Turner RS, Hebron ML, Lawler A, et al. Nilotinib Effects on Safety, Tolerability, and Biomarkers in Alzheimer’s Disease. Ann Neurol 2020;88(1):183–194.

33. Pagan FL, Hebron ML, Wilmarth B, et al. Nilotinib Effects on Safety, Tolerability, and Potential Biomarkers in Parkinson Disease: A Phase 2 Randomized Clinical Trial. JAMA Neurol 2020;77(3):309–317.

34. Simuni T, Fiske B, Merchant K, et al. Efficacy of Nilotinib in Patients With Moderately Advanced Parkinson Disease: A Randomized Clinical Trial. JAMA Neurol 2020.

35. Dobson-Stone C, Velayos-Baeza A, Filippone LA, et al. Chorein detection for the diagnosis of chorea-acanthocytosis. Annals of neurology 2004;56(2):299–302.

36. Kuhle J, Plattner K, Bestwick JP, et al. A comparative study of CSF neurofilament light and heavy chain protein in MS. Mult Scler 2013;19(12):1597–1603.

37. Keshavan A, Heslegrave A, Zetterberg H, Schott JM. Stability of blood-based biomarkers of Alzheimer’s disease over multiple freeze-thaw cycles. Alzheimers Dement (Amst) 2018;10:448–451.

38. Beste C, Stock AK, Zink N, Ocklenburg S, Akgun K, Ziemssen T. How minimal variations in neuronal cytoskeletal integrity modulate cognitive control. NeuroImage 2019;185:129–139.

39. Akgün K, Kretschmann N, Haase R, et al. Profiling individual clinical responses by high-frequency serum neurofilament assessment in MS. Neurology - Neuroimmunology Neuroinflammation 2019;6(3):e555.

40. Matte A, De Falco L, Federti E, et al. Peroxiredoxin-2: A Novel Regulator of Iron Homeostasis in Ineffective Erythropoiesis. Antioxid Redox Signal 2018;28(1):1–14.

41. Beneduce E, Matte A, De Falco L, et al. Fyn kinase is a novel modulator of erythropoietin signaling and stress erythropoiesis. Am J Hematol 2019;94(1):10–20.

42. Honisch S, Gu S, Vom Hagen JM, et al. Chorein Sensitive Arrangement of Cytoskeletal Architecture. Cellular physiology and biochemistry : international journal of experimental cellular physiology, biochemistry, and pharmacology 2015;37(1):399–408.

43. Olivieri O, De Franceschi L, Bordin L, et al. Increased membrane protein phosphorylation and anion transport activity in chorea-acanthocytosis. Haematologica 1997;82(6):648–653.

44. De Franceschi L, Fumagalli L, Olivieri O, Corrocher R, Lowell CA, Berton G. Deficiency of Src family kinases Fgr and Hck results in activation of erythrocyte K/Cl cotransport. J Clin Invest 1997;99(2):220–227.

45. Steegmann JL, Baccarani M, Breccia M, et al. European LeukemiaNet recommendations for the management and avoidance of adverse events of treatment in chronic myeloid leukaemia. Leukemia 2016;30(8):1648–1671.

46. Khalil M, Teunissen CE, Otto M, et al. Neurofilaments as biomarkers in neurological disorders. Nature reviews Neurology 2018;14(10):577–589.

